# Control Over Practice as a Mediator Between Role Overload and Job Strain Among Hospital Nurses: A Structural Equation Modeling Approach

**DOI:** 10.64898/2025.12.05.25341300

**Authors:** Emad Adel Shdaifat

## Abstract

**Objective:** This study aimed to investigate whether control over practice plays a mediating role in the relationship between job strain and role overload among nurses in a hospital setting.

**Methods:** A cross-sectional survey was conducted among 306 nurses selected using a stratified random sampling technique. The Working Condition Scale was used to collect demographic information and assess working conditions, while job strain was measured using nine statements across the four dimensions. The data were analyzed using Smart PLS 3.

**Results:** The structural model was evaluated by examining the R^2^, Q^2^, and significance of the paths. Hypothesis testing revealed significant associations between role overload and job strain (β = 0.904, t = 7.072, p = 0.000), role overload and control over practice (β = -0.649, t = 11.045, p = 0.000), and control over practice and job strain (β = 0.259, t = 1.976, p = 0.048). The mediation analysis showed that control over practice fully mediated this relationship (β= -0.168, t = 1.766, p = 0.078).

**Conclusion:** Enhancing control over work can alleviate adverse effects of role overload on job strain among nurses. This finding has crucial implications for organizations to reduce job strain. Future studies should explore alternative strategies.

**Summary statement:** *What is already known about this topic?:*

- Role overload is recognized as a significant contributor to job strain among nurses, particularly in high-pressure hospital environments.
- Control over practice has been proposed as a potential mitigating factor; however, its specific mediating role remains inadequately explored.

*What this paper adds?:*

- This study presents empirical evidence demonstrating that control over practice significantly mediates the relationship between role overload and job strain.
- It emphasizes that increased control over practice among nurses mitigates the adverse effects of role overload on job strain.
- Structural Equation Modeling substantiates the presence of a full mediation effect, thereby enhancing the understanding of the underlying mechanisms involved.

*The implications of this paper:*

- Strengthening nurses’ autonomy in their professional roles represents a strategic intervention aimed at alleviating job strain and enhancing overall well-being.
- It is imperative for hospitals and healthcare policymakers to prioritize organizational reforms that empower nursing personnel within their professional environments.
- The findings of this study advocate for the formulation of targeted programs and leadership practices that foster autonomy and encourage participatory decision-making among nursing staff.

## Introduction

Job strain is a prevalent issue among healthcare workers, including nurses, and can lead to negative outcomes, such as burnout, decreased job satisfaction, and higher rates of turnover (Adriaenssens et al., 2015; Shanafelt et al., 2012). Role overload, defined as the perception of having too many demands and insufficient resources to meet them, is a key factor contributing to job strain among nurses (Bakker et al., 2014). Recent research indicates that granting nurses control over their practice, which refers to their level of independence in their work, can alleviate the adverse consequences of job demands on job strain (Hunsaker et al., 2015; Spence Laschinger et al., 2016).

Although literature has established the individual effects of role overload and control over practice on job strain, little is known about the interplay between these two factors and how they collectively influence job strain among nurses. Moreover, it is unclear whether control over practice serves as a mediator or moderator in the relationship between role overload and job strain. Investigating these relationships is important because they can provide insights into potential interventions or strategies to help nurses better manage their job demands and maintain their well-being.

Several recent studies support the hypothesis that control over practice is an important factor in reducing job strain and promoting nurses” wellbeing. For example, 5 found that nurses with greater autonomy in their work reported lower levels of emotional exhaustion and depersonalization, which are fundamental elements of burnout. Similarly, 4 found a correlation between higher levels of control over practice and the reduced levels of job strain experienced by nurses.

Recent studies have proposed that control over practice can act as a mediator in the association between role overload and job strain. 6 found that control over practice partially mediates the relationship between workload and job strain among Chinese nurses. Similarly, 7 discovered that control over practice played a partial mediating role in the correlation between role ambiguity and job satisfaction among Korean nurses.

Investigating the interplay between role overload and control over practice, and the potential mediating role of control over practice in the relationship between role overload and job strain, is important because it can inform interventions and strategies to help nurses better manage job demands and maintain their well-being. For example, interventions aimed at increasing nurses” control over their work, such as providing opportunities for professional development and training, may be effective in reducing job strains and promoting well-being. Additionally, interventions aimed at reducing role overload, such as workload management and staffing, may be effective in improving nurses” well-being and quality of patient care.

Furthermore, a study by 8 found that role overload and lack of control over practice were significant predictors of burnout among nurses. This study also revealed that control over practice had a moderating effect on the correlation between role overload and burnout. This indicates that nurses with more autonomy at work experience less burnout, even in situations where they have high levels of role overload. This result supports the idea that control over practice can function as a protective factor against the adverse impacts of job demands on job strain.

Additionally, a meta-analysis by 9 examined the relationship between job demands, control, and burnout among healthcare workers including nurses. A meta-analysis found that both job demands and controls were significant predictors of burnout, with job demands having a stronger effect. Nevertheless, this study also revealed that control over practice moderates the association between job demands and burnout. This implies that nurses with more independence in their work experience lower levels of burnout even when facing high levels of job demands.

These studies further support the importance of investigating the interplay between role overload and control over practice in relation to job strain among nurses. Overall, the results suggest that control over practice can safeguard against the detrimental impact of job demands on job strain, including burnout, among nurses.

Moreover, research has shown that interventions targeting control over practice can be effective in reducing job strain and improving nurses” wellbeing. For example, 10 implemented a job crafting intervention among nurses that aimed to increase control over their work. The intervention involved providing nurses with the tools and resources to redesign their job tasks and roles to better fit their strengths and interests. The study found that job crafting interventions led to reductions in job strain and burnout among nurses as well as increased job satisfaction and engagement.

Another study by 11 implemented an autonomy-supportive leadership intervention among nurses that aimed to increase their control over their work. The intervention involved providing nurses with support and resources to make decisions about their work as well as recognition and feedback for their achievements. The study found that autonomy-supportive leadership interventions led to reductions in job strain and burnout among nurses as well as increased job satisfaction and engagement.

Overall, the literature suggests that control over practice is a crucial factor in mitigating job strain among nurses and that interventions targeting control over practice can be effective in reducing job strain and improving well-being among nurses. Investigating the interplay between role overload and control over practice in relation to job strain among nurses can provide valuable insights into potential interventions or strategies that can be implemented to help nurses better manage job demands and maintain their well-being, ultimately improving patient care quality and reducing health care costs.

This study aimed to investigate the protective function of control over practice in mitigating the unfavorable effects of role overload on job strain in a hospital setting. We hypothesized that nurses with greater autonomy in their work would report reduced levels of job strain, even in situations where they experienced high levels of role overload. Additionally, this study proposes that control over practice acts as a mediator in the correlation between role overload and job strain. This suggests that nurses with higher levels of control over their work experience lower levels of job strain even when they experience high levels of role overload. The study is guided by the following hypotheses (Figure 1):

**H1:** role overload significantly affects job strain.

**H2:** role overload has a significant effect on control over practices.

**H3:** control over practices has a significant effect on job strain.

**H4:** Control over practice mediates the relationship between role overload and job strain

**Figure 1.**
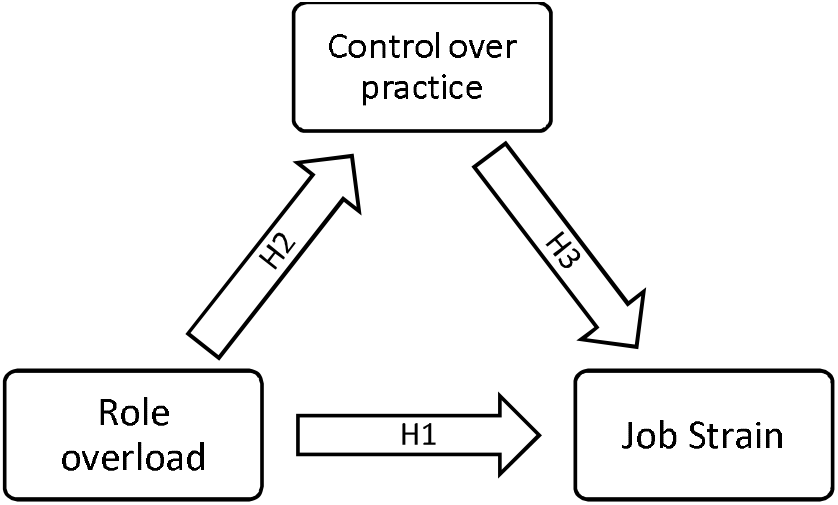
Conceptual Model

## Methods

### Setting and design

This study employed a descriptive cross-sectional survey to examine the demographics and working conditions of nurses within a teaching hospital. The survey was conducted within a teaching hospital and the participants were nurses in the facility. The data for this study were collected as part of a larger project titled “Absenteeism Among Nurses: Costs, Working Conditions, and Related Factors Study” (Shdaifat et al., 2021).

### Sample size and sample technique

The Raosoft sample size calculator (Raosoft, 2004) was used to determine the sample size based on specific parameters. These parameters included a margin of error of 5%, confidence level of 95%, population size of 730 nurses, and response distribution of 50%. The minimum required sample size was 265. Eligible participants were selected based on specific inclusion criteria designed to align with the research objectives and ensure a representative sample. Eligible participants were registered nurses currently employed in hospitals who were actively involved in clinical roles. To ensure familiarity with the hospital environment, participants were required to have at least six months of experience in their current hospital. Additionally, they were required to be proficient in English and provided informed consent to participate. The exclusion criteria were nurses in non-clinical roles, such as those primarily performing administrative, managerial, or educational tasks without direct patient care responsibilities. Intern nurses were excluded from this study. A stratified random sampling technique was employed to select nurses eligible to participate in the study. The selection process was based on the units, number of nurses, and the type of shift to which each nurse was assigned. This approach ensured that the sample was representative and that the data collected from nurses were applicable to the entire population of nurses in the hospital. The final sample consisted of 306 participants included in the analysis.

### Study tool

Data were collected using self-administered questionnaires, which included demographic questions and validated scales for assessing working conditions. The Working Condition Scale used in this study was adapted from the Canadian National Survey of the Work and Health of Nurses conducted in 2005 by Shields and Wilkins (Shields & Wilkins, 2006). This scale uses a Likert-type format with five response options ranging from strongly disagree to strongly agree. Scores were calculated for each of the following subscales: role overload, which included five items, and control over practice, which included seven items. Each subscale includes a set of statements, with scores ranging from 0 to a certain value, with higher scores indicating higher levels of the measured construct. Job strain was evaluated using nine statements across three dimensions: psychological demand, skill discretion, and decision authority. The highest score for each dimension indicates higher job strain. Cronbach”s alpha for the control over practice scale was 0.91, while that for the role overload scale was 0.84 (Shields & Wilkins, 2006). Regarding job strain, Cronbach”s alpha for job strain exceeded 0.70, indicating acceptable internal consistency (Karasek et al., 1998).

### Ethical consideration

This study was conducted in accordance with the ethical standards set forth in the Declaration of Helsinki, which offers guidelines for research involving human participants. The study received ethical approval from the Institutional Review Board (IRB) at Imam Abdulrahman Bin Faisal University in Saudi Arabia (IRB-2017-04-159). Prior to participation, potential participants were provided with a comprehensive information sheet explaining the research objectives, significance, and potential benefits, and verbal explanations were provided to ensure clear understanding. Participation was voluntary, and written consent was obtained from each participant. The study strictly followed data protection regulations and institutional policies to protect the participants” confidentiality and anonymity. Identifying information, including the name of the study setting, was securely safeguarded and not disclosed.

### Data Analysis

Data were entered and cleaned in Microsoft Excel before being analyzed using SmartPLS 3. Convergent validity was verified by examining factor loadings, composite reliability, and average variance extracted, through the employment of the SmartPLS 3 software. Prior to conducting the structural equation modeling (SEM) analysis, normality and multicollinearity assumptions were evaluated. Categorical variables were analyzed using frequencies and percentages, whereas continuous variables were expressed as means and standard deviations. The relationship between role overload, control over practice, and job strain was determined using Smart PLS with latent variables for each construct in the model and path coefficients calculated for each variable. Bootstrap resampling with 5000 subsamples was used to test the significance of each path coefficient.

## Results

### Participant characteristics

The study participants were distinguished by a range of demographic factors. The mean age of the participants was 35.4 years, and the average duration of their nursing career was 11.0 years. The majority of the participants were female (89.5%), married (66.0%), and held a bachelor”s degree (74.5%). In terms of employment, over half of the participants worked in critical units (52.0%), primarily during the A shift (46.1%). Additionally, a large majority of the nurses reported being free of chronic diseases (93.1%) and having children (57.2%).

### Measurement Model

This study used Cronbach”s alpha and composite reliability (CR) to evaluate the reliability of the constructs. Table 1 presents all CRs. It is generally considered acceptable for the value of Cronbach”s alpha to fall within the range 0.581–0.797 (Shi et al., 2012). This study evaluated the convergent validity of the constructs using Average Variance Extracted (AVE), which ranged from 0.317 to 0.613. According to (Fornell & Larcker, 1981), a construct”s convergent validity is considered satisfactory if the AVE is less than 0.5, but composite reliability is greater than 0.6. The factor loadings of the study, which are presented below, met the minimum threshold for structural interpretation, with a range of 0.30 to 0.40, as per (J. Hair et al., 2006). All factors and their subscales were statistically significant. Table 1 contains the results for reliability and validity as well as the factor loadings for the items.

**Table 1.**
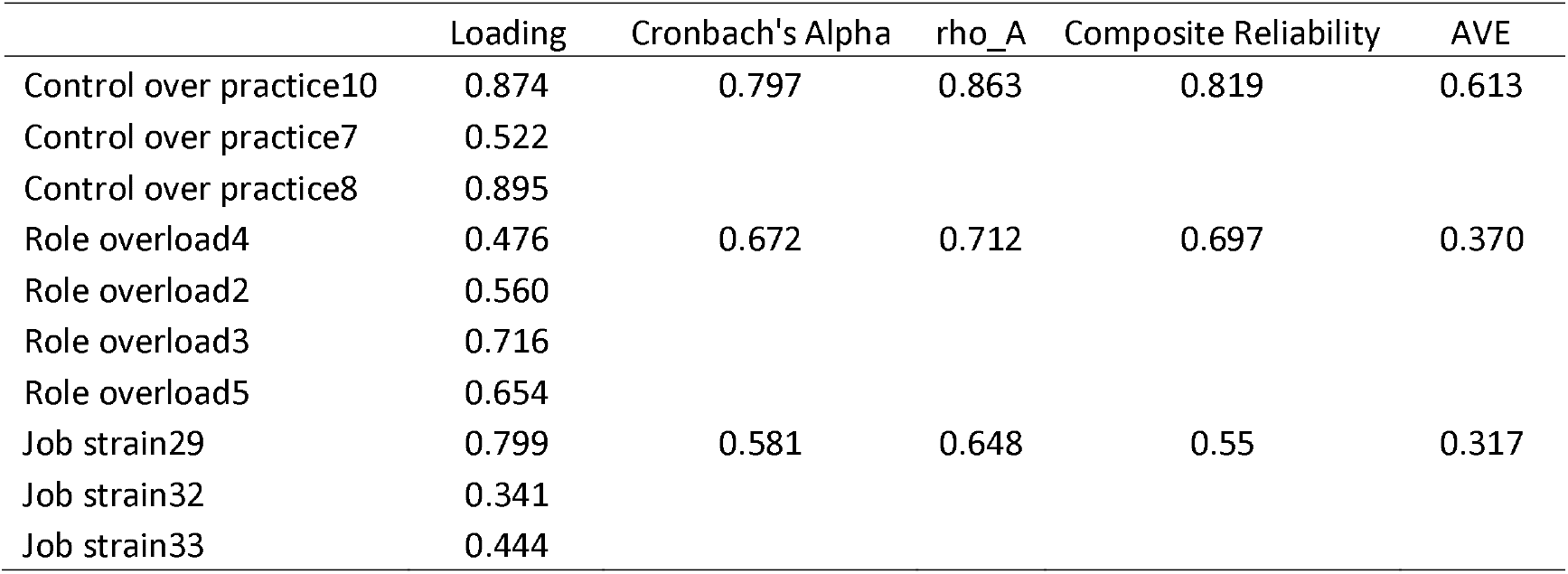
Factor Loading, Cronbach alpha, rho A, composite reliability and AVE.

Discriminant validity was assessed using the Fornell-Larcker criterion, which revealed that the square root of the AVE for each construct was greater than the inter-construct correlation (Table 2). The heterotrait-monotrait ratio of correlations was also employed to evaluate discriminant validity, with values below the threshold of 0.90. The findings indicate that the study had established discriminant validity (Table 3).

**Table 2.**
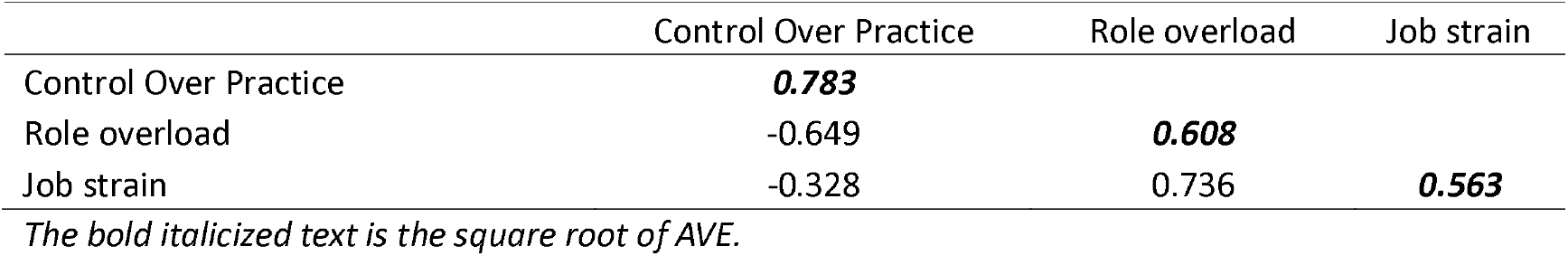
Fornell-Larcker Criterion discriminant validity.

**Table 3.** Heterotrait-Monotrait Ratio of Correlations (HTMT) discrimination validity of the measurement model.

|  | Control Over Practice | Role overload |
| --- | --- | --- |
| Role overload | 0.697 | - |
| Job strain | 0.304 | 0.693 |

### Structure Model

The structural model represents the anticipated connections between the variables in the research framework. To assess the structural model, R^2^, Q^2^, and the significance of the paths were evaluated. This study employed 5000 resamples to produce 95% confidence intervals, which are presented in Table 4. A confidence interval that deviates from zero indicates a significant relationship. The findings of the hypothesis testing are summarized in Table 4.

**Table 4.** Results of hypothesis testing, R^2^, Q^2^ and SRMR.

| H | Paths | β | SD | T Statistics | P Values |  |
| --- | --- | --- | --- | --- | --- | --- |
| H1 | Role overload -> Job strain | 0.904 | 0.128 | 7.072 | 0.000 |  |
| H2 | Role overload -> Control over practice | -0.649 | 0.059 | 11.045 | 0.000 |  |
| H3 | Control over practice -> Job strain | 0.259 | 0.131 | 1.976 | 0.048 |  |
| Construct |  | R<sup>2</sup> |  | Q<sup>2</sup> |  |  |
| Control over practice |  | 0.421 |  | 0.179 |  |  |
| Job strain |  | 0.581 |  | 0.108 |  |  |
| Model |  | SRMR | d_ ULS | d_ G | Chi-Square | NFI |
| Saturated Model |  | 0.086 | 0.409 | 0.117 | 184.062 | 0.819 |
| Estimated Model |  | 0.086 | 0.409 | 0.117 | 184.062 | 0.819 |

The quality of the structural model was assessed using the R^2^ value, which measured the strength of each structural path for the dependent variable (Juan et al., 2017). An R2 value equal to or greater than 0.1 (Falk & Miller, 1992) is deemed adequate. Based on the results displayed in Table 4, all the R^2^ values surpassed 0.1, indicating the robust predictive capacity of the model. The Q^2^ values also indicated that the model had predictive relevance, with all constructs exhibiting significance (Table 4). Moreover, the SRMR value of 0.08 suggests an acceptable model fit (J. F. Hair et al., 2017).

Various hypotheses were tested to determine the significance of the relationships and to assess the model”s goodness of fit. H1 investigated whether there is a significant relationship between role overload and job strain. The results indicated that role overload had a significant impact on job strain (β = 0.904, t = 7.072, p = 0.000), thus supporting H1. Similarly, H2 reveals that role overload has a significant impact on control over practice (β = - 0.649, t = 11.045, p = 0.000), which also received support. Finally, H3 investigated whether control over practice has a significant impact on job strain. The results demonstrated a significant impact (β = 0.259, t = 1.976, p = 0.048), thus supporting H3.

### Mediation Analysis

Mediation analysis was performed to assess the mediating role of control over practice in relation to Hypothesis 4. The results (Table 5) reveal the full mediating role of control over practice (β= -0.168, t = 1.766, p = 0.078). Consistent with previous research, indirect effects can occur without fulfilling the criteria of the causal-stage approach. Bootstrapping analysis can determine the significance of the product of DV and IV, which indicates the significance of the mediation model or indirect effect (Gürbüz & Bayık, 2021).

**Table 5.**
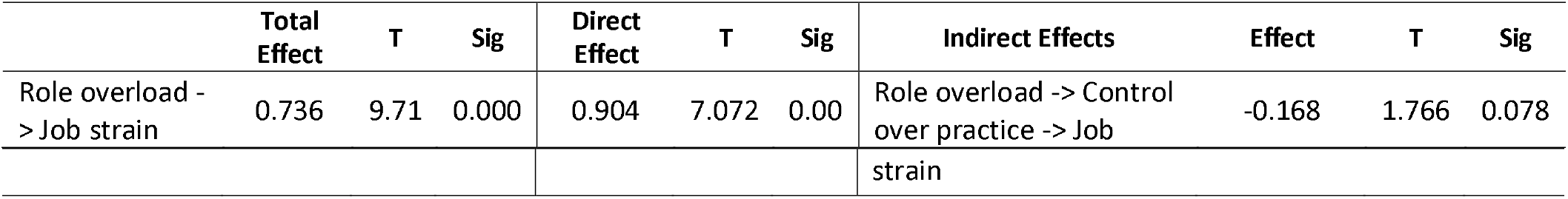
Total, Direct, Specific Indirect Effects.

## Discussion

A mediation analysis was conducted to examine the role of control over practice in job strain. The results indicated full mediation, suggesting that control over practice significantly mediates the relationship between role overload and job strain. Bootstrapping analysis confirmed the significance of indirect effects. Hypothesis testing showed that role overload has a significant impact on job strain and control over practice. Moreover, control over practice had a significant impact on job strain. Overall, the results support the hypotheses of this study, indicating that control over practice plays a significant mediating role in reducing job strain caused by role overload.

This study”s finding that role overload has a significant effect on job strain is in line with previous research. For instance, a study by (Demerouti et al., 2001) found that role overload was positively associated with emotional exhaustion and depersonalization among healthcare workers. Similarly, a meta-analysis conducted by (Bakker et al., 2004) established that role overload was positively associated with burnout.

These results are consistent with previous research that has identified role overload and lack of control over work as important predictors of job strain (Schaufeli & Bakker, 2004). The finding that control over practice mediates the relationship between role overload and job strain is particularly noteworthy, because it suggests that providing employees with more control over their work can mitigate the negative effects of role overload on job strain. This study”s finding that control over practice has a significant impact on job strain is also in line with previous research. For instance, a study by (Hakanen et al., 2006) found that job control, which encompasses control over practice, was negatively related to burnout among healthcare workers. Similarly, a meta-analysis conducted by (Halbesleben & Buckley, 2004) established that job control is negatively associated with emotional exhaustion and depersonalization.

The finding that control over practice mediates the relationship between role overload and job strain is also consistent with previous research. For example, a study by (Xanthopoulou et al., 2007) found that job resources (including control over practice) mediated the relationship between job demands and burnout among healthcare workers. Similarly, a meta-analysis by (Halbesleben & Buckley, 2004) found that job control partially mediated the relationship between job demands and emotional exhaustion.

This study has several implications for organizations. First, organizations should take steps to reduce role overload by providing employees with the necessary resources to manage their workload effectively. This can include providing additional staff, training, or technology to help employees prioritize their tasks. Second, organizations should provide employees with more control over their work by offering autonomy and decision-making authority. This can help employees manage their workloads and reduce the negative impact of role overload on their job strain.

However, it is important to note that this study had some limitations. For example, the study was conducted in a specific setting (e.g., a hospital), which may limit the generalizability of the results. Additionally, the study used self-reported measures, which may be subject to bias.

In summary, this study”s conclusion that control over practice has a significant mediating role in reducing job strain resulting from role overload has vital implications for organizations. Providing employees more control over their work can help alleviate the adverse effects of role overload on job strain. In the future, further techniques should be investigated to reduce job strain and to identify the underlying mechanisms of the connection between role overload, control over practice, and job strain.

## Limitation of the study

One limitation of this study was the use of self-reported measures, which can introduce response bias and limit the accuracy of the data. Another limitation is the study”s narrow focus on nurses in a hospital setting, which may restrict its generalizability to other healthcare contexts. In addition, the study”s cross-sectional design prevented the establishment of causal relationships between variables. The sample size and sampling technique could also affect the representativeness of the findings, and the use of only one measurement tool limited the comprehensiveness of the assessment.

## Implications

The findings of this study have significant implications for organizations seeking to address job strain among nurses. By emphasizing the crucial role of practice control in mitigating the negative effects of role overload on job strain, organizations can implement interventions that prioritize granting nurses greater autonomy and decision-making power. Strategies such as workload management, interventions to adjust staffing levels, and opportunities for professional development can empower nurses to effectively handle their work demands and improve their overall well-being. Furthermore, this study highlights the importance of addressing role overload and promoting practice control to enhance job satisfaction, decrease burnout, and enhance the quality of patient care.

## Conclusion

The study concluded that control over practice is important for reducing job strain among nurses, especially in cases of role overload. By giving nurses more autonomy and decision-making power, organizations can effectively lessen the negative effects of role overload on job strain, leading to improved well-being and job satisfaction. These findings highlight the significance of addressing role overload and promoting control over practice as key strategies for enhancing nurses” work conditions, and ultimately improving the quality of patient care in healthcare settings.

## Data Availability

All data produced in the present study are available upon reasonable request to the authors

## Notes

### Competing Interest Statement

The authors have declared no competing interest.

### Funding Statement

This study did not receive any funding

### Author Declarations

The data for this study were collected through an online survey administered to eligible participants. The study received ethical approval from the Institutional Review Board (IRB) at Imam Abdulrahman Bin Faisal University in Saudi Arabia (IRB-2017-04-159).

